# Ensemble Approach for Predicting the Diagnosis of Osteoarthritis Using Soft Voting Classifier

**DOI:** 10.1101/2023.01.27.23284757

**Authors:** Jun-hee Kim

**Affiliations:** Sleep&Spine, Research & Development, KOREATECH Corporation

**Author notes:** **Corresponding author:** Jun-hee Kim, PhD, PT, Senior Researcher, Address: 12 Bongeunsa-ro 49-gil, Gangnam-gu, Seoul, Republic of Korea (06103), Spine&Sleep, Research & Development, KOREATECH Corporation.

## Abstract

**Background:** Osteoarthritis (OA) is a common degenerative disease of the joints. Risk factors for OA include non-modifiable factors such as age and gender and modifiable factors such as physical activity.

**Purpose:** This study aimed to construct a soft voting ensemble model to predict OA diagnosis using variables related to individual characteristics and physical activity and to identify important variables in constructing the model through permutation importance.

**Method:** Using the RFECV technique, the variables with the best predictive performance were selected among variables, and an ensemble model combining the RandomForest, XGBoost, and LightGBM algorithms was constructed, and the predictive performance and permutation importance of each variable were evaluated.

**Result:** The variables selected to construct the model were age, gender, grip strength, and quality of life, and the accuracy of the ensemble model was 0.828. The most important variable in constructing the model was age (0.199), followed by grip strength (0.053), quality of life (0.043), and gender (0.034).

**Conclusion:** The performance of the model for predicting OA was relatively good, and if this model is continuously used and updated, this model could readily be used to predict OA diagnosis and the predictive performance of OA may be further improved.

## Introduction

Osteoarthritis (OA) is a common degenerative disease of joints and surrounding structures, and it is estimated that there approximately 300 million patients worldwide suffer from OA.^1^ Additionally, due to medical expenses and income loss due to OA, social losses of more than 303 billion dollars are generated annually in the United States.^2^ For individuals, pain or loss of function due to OA can reduce activities of daily living, which can limit social participation as well as reduce quality of life.^3^

Since OA has such a high prevalence, many studies have been conducted on risk factors related to OA. Risk factors associated with OA, like other diseases, include modifiable and unmodifiable factors.^4^ Non-modifiable risk factors associated with developing OA include age and gender.^4^ Aging is considered the greatest risk factor for developing OA.^5^ The incidence of OA begins to increase rapidly from the age of 50 and stops or decreases after the age of 70.^6^ Gender is also a nonmodifiable factor in the incidence of OA, and the relative risk of hand, knee, and hip OA was found to be 1.52 times greater in women than in men.^7^ However, this tendency may differ with age. After the age of 50, the risk of developing OA is higher in women than in men, but OA may be more common in men than in women under the age of 50.^8^

Body composition, including height and weight, and physical activity factors, including occupational and recreational activities, can be viewed as modifiable risk factors for developing OA.^4^ Obesity can be seen as a risk factor for developing OA, and the risk of developing OA in obese or overweight people was 2.96 times higher than in people with normal weight.^9^ Additionally, occupational groups with excessive physical activity, such as frequent sitting and standing up, frequent use of joints, or repetitive lifting of objects, have been reported to have a high risk of OA.^10,11^ When repetitive loads are applied to joints by participation in leisure activities such as sports, OA can occur even in young people due to damage to joints and surrounding structures.^12^ In particular, those who had experience participating in sports with a high rate of physical contact and high trauma potential, such as soccer, showed a higher rate of OA when they became older.^12,13^

Machine learning is a technique of predicting new data based on attributes learned from a large amount of data.^14^ Machine learning shows great performance in estimating values and predicting classifications based on training data.^14^ Recently, studies using machine learning to predict the risk of disease or to increase the accuracy of diagnosis using large amounts of medical data are actively being conducted in the medical field.^15^ With the development of machine learning techniques in the 2000s, models using algorithms such as support vector machine (SVM) and principal component analysis (PCA) have begun to be applied to knee OA research.^16^ These studies mainly used medical images such as X-ray or MRI, or biomechanical data such as kinematics including joint angles or accelerations.^16^ Various algorithms are developed and used to build machine learning models, and new algorithms are continuously updated. For example, the XGBoost developed by Chen (2015) and the LightGBM algorithm proposed by Ke (2017) have been developed and actively used as boosting techniques to exhibit higher performance.^17,18^ Additionally, machine learning can be implemented by combining the previously developed techniques, such as ensembles, to construct new models and use them to predict data.^19^ The soft voting ensemble technique is a model merging technique that makes predictions through majority voting based on the prediction probabilities of different machine learning models.^20^ The soft voting ensemble technique can provide better overall results than other base models.^20^

Although a machine learning model that predicts the presence of OA in a specific joint using X-ray or MRI image or biochemical data has been created, a predictive model for OA diagnosis using general information such as individual characteristics including age, gender, height, and weight, and physical activity-related variables including occupational status and leisure activities has not been studied. Therefore, this study aimed to construct a soft voting ensemble model to predict OA diagnosis by using data such as age, gender, individual characteristics, and physical activities of Koreans in the KNHANES (The Korea National Health and Nutrition Examination Survey) database. Additionally, based on the constructed ensemble model, permutation importance is analyzed to identify variables with high importance in predicting OA diagnosis.

## Methods

### 1. Study design and population

The study samples were obtained from the KNHANES, a national periodic statistic conducted by the Korea Disease Control and Prevention Agency. This study used data from the first year of the 8th KNHANES (2019-2020). Among the 8110 datasets, only samples aged 19 years or older were selected. A total of 621 samples diagnosed with OA but not diagnosed with rheumatoid arthritis and 4988 samples with neither diagnosed OA nor rheumatoid arthritis were used to construct the model.

### 2. Variable selection

The variables used in this study, variables representing individual characteristics and variables related to physical activity were selected. The variables and explanations selected in this study are shown in Table 1.

**Table 1.**
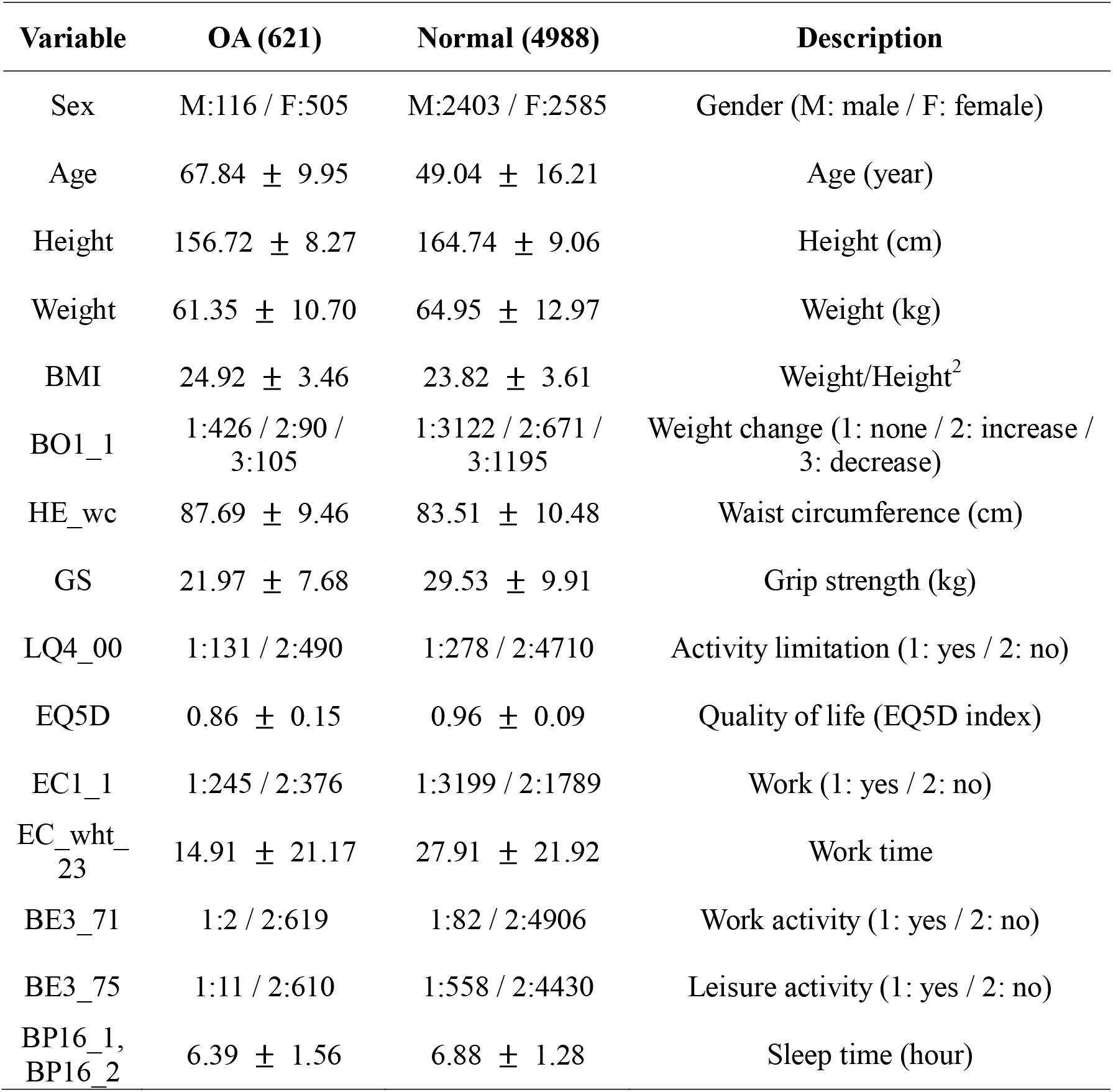
List of variables and variable descriptions

### 3. Data preprocessing

Boxplot was used to remove outliers from numerical data based on the interquartile range (IQR). The ratio of the number of samples of patients with OA to the number of samples of normal subjects was about 1:8, indicating an imbalance in the data distribution. To address the data imbalance, this study used the RandomUnderSampler function, which randomly selects the same number of normal samples as samples diagnosed with OA. Categorical variables were transformed into dummy and numerical variables were scaled with the StandardScaler function. The preprocessed data were classified into training and test data at a ratio of 7:3.

### 4. Feature selection

The machine learning algorithms used in this study were RandomForest, XGBoost, and LightGBM. In constructing these models, RFECV (Recursive Feature Elimination with Cross Validation) module was used to remove unnecessary variables. Since the same number of features was required to create an ensemble model by combining the three models, the features selected from the model with the best performance among the three models were used to construct the ensemble model.

### 5. Machine learning modeling

The data was divided into a training data set and a validation data set (hold-out validation), and models of the RandomForest, XGBoost, and LightGBM were built using the training data set, respectively. GridSearchCV was used to find the optimal hyperparameters for each model, and the optimal hyperparameters were set for each model. To improve the accuracy of OA diagnosis prediction, modeling was performed using the soft voting ensemble technique. In this study, an ensemble model was constructed with RandomForest, XGBoost, and LightGBM classifiers to predict OA diagnosis.

### 6. Evaluation

The performance of the model was evaluated by classifying the predicted and actual classes into a confusion matrix in the form of TP (true positive), FP (false positive), FN (false negative), and TN (true negative) using test data. Models were evaluated by accuracy, precision, recall, F1-score and ROC AUC score calculated based on the confusion matrix. The permutation importance was calculated to identify important variables in the model. Permutation importance indicates the relative importance of variables in construction a model.

## Results

### 1. Feature selection (RFECV)

As a result of performing the RFECV technique, the XGBoost model showed the best performance based on the smallest number of variables (Table 2). The features selected when constructing the ensemble model were Gender, Age, Quality of life, and Grip strength variables based on XGBoost, which showed the highest performance.

**Table 2.** Feature selection and mean test score of the three models

| Model | Feature selection (count) | Mean test score |
| --- | --- | --- |
| RandomForest | Gender, Age, Height, Weight, Waist circumference, BMI, Quality of life, Work time, Sleep time, Weight change, Grip strength (11) | 0.774 |
| XGBoost | Gender, Age, Quality of life, Grip strength (4) | 0.779 |
| LightGBM | Gender, Age, Height, Weight, Waist circumference, BMI, Quality of life, Work, Work time, Leisure activity, Sleep time, Weight change, Grip strength (13) | 0.756 |

### 2. Model evaluation

The confusion matrix of the ensemble model combining the three models using the validation data set is shown in Table 3. The accuracy score, an overall performance indicator calculated through the confusion matrix, was 0.828. The recall score, which indicates how well the model can identify actual positives, was 0.882. Additionally, the F-1 score was 0.837 and the ROC AUC score was 0.829.

**Table 3.**
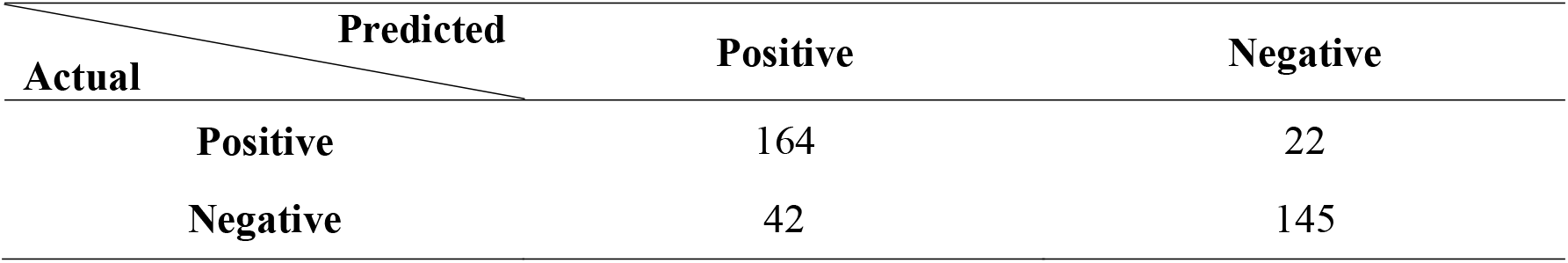
Confusion matrix of voting ensemble classification

### 3. Permutation Importance

Figure 2 shows the permutation importance of the ensemble model for predicting OA diagnosis. The most important variable in constructing the model was age, which was 0.199. The second important variable was grip strength, with a significance score of 0.053. The importance of quality of life was 0.043, and gender was 0.034.

**Figure 1.**
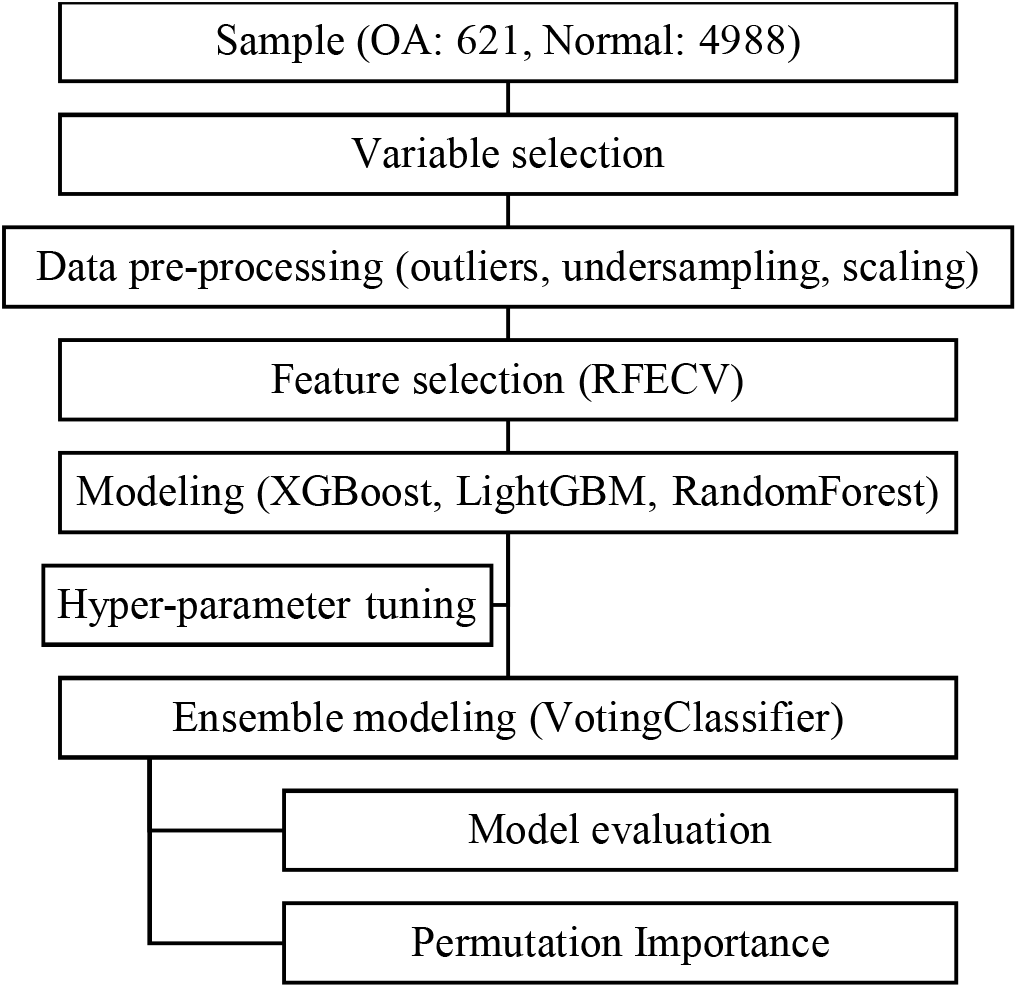
The flowchart diagram of the research process.

**Figure 2.**
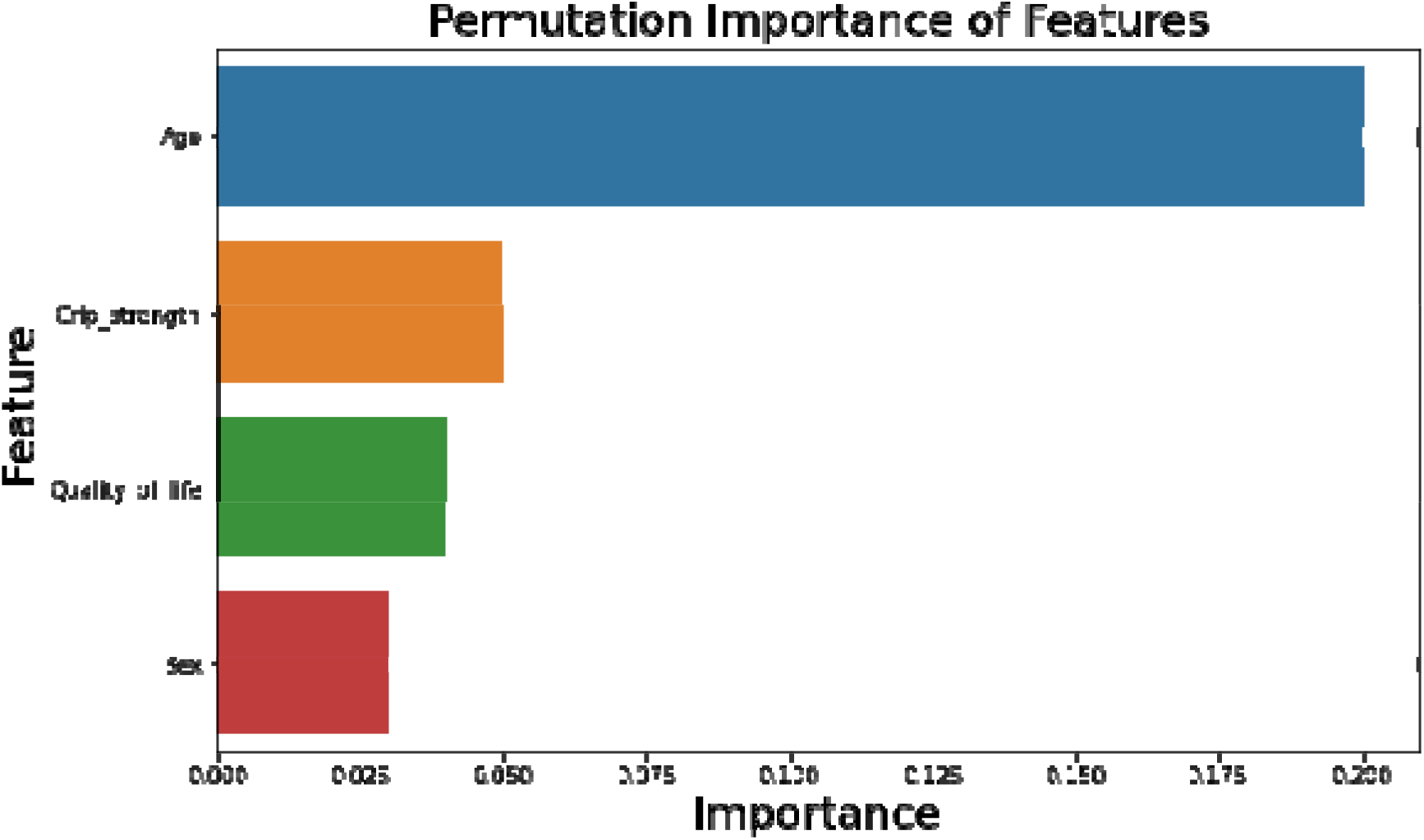
Permutation importance of features in the model

## Discussion

In this study, a machine learning classification model was constructed to predict OA diagnosis based on factors related to individual characteristics and physical activities during daily life. The variables selected to build the model were age, gender, grip strength, and quality of life. This classification model was created by combining the latest bagging and boosting classifiers such as RandomForest, XGBoost, and LightGBM. The classification model composed of these ensemble techniques had an accuracy of over 0.8 in predicting OA diagnosis. The variable with the highest importance in constructing the model was age, followed by grip strength, quality of life, and gender in order of importance.

In this study, the non-modifiable factors selected as variables in constructing the ensemble model were age and gender. OA is considered a degenerative joint disease, and its prevalence is reported to increase with age.^21–23^ Furthermore, studies have shown that age is the most prominent risk factor for the onset and progression of primary OA in joints such as the interphalangeal, hip and knee.^7,22,23^ OA can be pathologically characterized by irregularly distributed cartilage loss in areas of increased load or synovial inflammation.^24^ With aging, articular joints including cartilage, subchondral bone, muscle, soft tissue, synovial membrane, and synovial fluid may be deformed, resulting in OA.^25^ Based on these results, age would have shown the highest permutation importance in constructing an ensemble model in this study. It showed about 4-times higher permutation importance compared to other variables. Additionally, older age and female gender were risk factors for OA.^26^ Studies of OA and gender have reported a higher prevalence of OA in women than in men.^26,27^ The prevalence of symptomatic radiographic knee OA was 4.6% higher in women than men in one large cohort study, and 5.2% higher in women than men in another cohort study.^28,29^ Additionally, compared with men, women with OA have more severe radiological findings and symptoms.^27^ However, the explanation for the difference in OA incidence between men and women is still insufficient. It is also unclear whether this is due to differences in the roles of sex hormones and reproductive factors, or whether it is due to weight or obesity, which can be influenced by hormonal factors.^30,31^ Therefore, although the prevalence rate between men and women shows a difference, it may not be a variable that can completely distinguish the diagnosis of OA because the explanation for the difference is not sufficient.^30,31^ For this reason, the gender variable was selected to generate the ensemble model, but it showed the lowest permutation importance among the variables.

The modifiable factors selected as variables in constructing the ensemble model were grip strength and quality of life. Previous studies to identify the relationship between grip strength and OA have been continuously conducted.^5^ These studies used grip strength as a variable and found that people with OA of the hand mainly had reduced grip strength compared to the general population, and it was thought that the risk of OA could be predicted through grip strength.^32–34^ Based on these results, it was thought that improving grip strength through training would help improve OA symptoms.^35^ However, grip strength is not only a marker for OA but also a marker for general health, and since grip strength can vary according to gender or physical characteristics, it was considered difficult to determine OA of the hand only with grip strength.^32^ Therefore, grip strength was also a factor adopted as a variable in constructing the model, but it would have shown permutation importance similar to other variables except age. The EQ-5D used to measure quality of life in this study consists of 5 items that measure motor skills, self-management, activities of daily living, pain/discomfort, anxiety/depression in 3 levels and 1 item that measures health status in a visual analog scale.^36^ Compared to assessment tools that measure the health status of OA patients, such as the Western Ontario and MacMaster Universities (WOMAC) OA index, the EQ-5D showed overall good validity and reliability in assessing the health status of patients with OA.^37,38^ Also, the EQ-5D has fewer and simpler questions, so it may be more appropriate for elderly subjects.^38,39^ Therefore, the EQ-5D is used to evaluate the overall health status of relatively elderly patients with OA.^40,41^ For this reason, the quality of life variable evaluated by EQ-5D would have been selected in constructing the ensemble model. Similarly, since the quality of life can be influenced by various other factors such as psychology, physical condition, and social environment, it would have shown low permutation importance compared with other variables.

In the KNee OsteoArthritis Prediction challenge held in 2020 (KNOAP2020), various machine learning models for predicting knee OA have been submitted using X-ray and MRI image and clinical data such as age, BMI, affected side, and history of knee injury.^42^ The model that showed the highest ROC AUC score in the competition was a model that applied the logistic regression algorithm using X-ray and clinical data and scored 0.636.^42^ The model with the next highest ROC AUC score was an ensemble model using X-ray, MRI, and clinical data, with a score of 0.624.^42^ According to Hosnijeh (2018), the ROC AUC score of the hip OA model, which was constructed by combining imaging, biochemical, and genetic variables with variables such as age, obesity, waist circumference, and gender, was between 0.67 and 0.82.^43^ Although image data such as X-ray or MRI and biomechanical or genetic factors were not used as variables, the ROC AUC score of the ensemble model constructed in this study was relatively high. The reasons for the relatively high performance of the ensemble model in this study may be the following difference. The data used in KNAOP2020 were obtained from the Osteoarthritis Initiative (OAI) database.^42^ The average age of the subjects in this dataset was over 61 years old.^42^ Additionally, in the Hosnijeh (2018) study, all of the selected subjects were over 55 years of age, and the average age of the samples used to construct the model ranged from 55.8 to 64 years^43^ The age of the dataset used in our study was over 19 years old, and the average age of subjects with OA was 67 years old, and the average age of normal subjects was 49 years old, showing an age difference of about 18 years. Additionally, while other studies have constructed prediction models for knee, hip, and wrist OA separately, this study constructed a model that included all OA without distinction. In the case of older subjects, the possibility of having OA in one of the body parts would be higher than that of younger subjects.^5,7,22^ Therefore, the age variable in the ensemble model constructed in this study showed great permutation importance in the classification compared with other variables, and it would have shown high classification performance despite the absence of imaging data, genetic, and biochemical data.

This study had several limitations. First, the constructed model showed relatively high predictive performance, but the ensemble model based on the machine learning algorithm cannot explain the causal relationship between OA and other variables. Second, because it is not a predictive model for OA of a specific joint, this model can predict the overall risk of OA, but it will be difficult to use to predict or diagnose OA of a specific joint. Based on these limitations, future research will require the development and application of an algorithm that can infer causal relationships, and modeling to predict arthritis of a specific joint will be needed.

## Conclusion

In this study, a model to predict OA was created using individual characteristics and physical activity-related variables. This model was constructed by selecting variables of age, gender, grip strength, and quality of life, and was composed of an ensemble model combining RandomForest, XGBoost, and LightGBM models with high predictive performance. The performance of the ensemble model for predicting OA diagnosis was relatively good, and this ensemble model can be used to easily predict whether a person will be diagnosed with OA. Additionally, if the model is updated by continuously accumulating new data, the prediction performance can be further improved.

## Data Availability

https://knhanes.kdca.go.kr/knhanes/eng/index.do

## Acknowledgments

This research did not receive any specific grant from funding agencies in the public, commercial, or not-for-profit sectors. This study was conducted using de-identified data from the 8th KNHANES performed by the Korea Disease Control and Prevention Agency in 2019-2020.

